# Normative values for body surface gastric mapping evaluations of gastric motility using Gastric Alimetry: spectral analysis

**DOI:** 10.1101/2022.07.25.22278036

**Authors:** Chris Varghese, Gabriel Schamberg, Stefan Calder, Stephen Waite, Daniel Carson, Daphne Foong, William Wang, Vincent Ho, Jonathan Woodhead, Charlotte Daker, William Xu, Peng Du, Thomas L Abell, Henry P. Parkman, Jan Tack, Christopher N. Andrews, Gregory O’Grady, Armen A Gharibans

**Author notes:** **Corresponding Author** Professor Greg O’Grady, Department of Surgery, University of Auckland, New Zealand,. Joint first authors (Chris Varghese, Gabriel Schamberg).

## Abstract

**Introduction:** Body surface gastric mapping (BSGM) is a new non-invasive test of gastric function. BSGM offers several novel and improved biomarkers of gastric function capable of differentiating patients with overlapping symptom-profiles. The aim of this study was to define normative reference intervals for BSGM spectral metrics in a population of healthy controls.

**Methods:** BSGM was performed in healthy controls using Gastric Alimetry (Alimetry, New Zealand) comprising a stretchable high-resolution array (8×8 electrodes; 196 cm^2^), wearable Reader, and validated symptom-logging App. The evaluation encompassed a fasting baseline (30 min), 482 kCal meal, and 4-hr postprandial recording. Normative reference intervals were calculated for BSGM metrics including the Principal Gastric Frequency, Gastric Alimetry Rhythm Index (GA-RI; a measure of the concentration of power in the gastric frequency band over time), BMI-adjusted amplitude (µV), and fed:fasted amplitude ratio (ff-AR). Data are reported as median and reference interval (5^th^ and/or 95^th^ percentiles).

**Results:** 110 subjects (55% female, median age 32 (IQR 24 – 50), median BMI 23.8 kg/m^2^ (IQR 21.4 – 26.9)) were included. The median Principal Gastric Frequency was 3.04 cpm; reference interval: 2.65 – 3.35 cpm. Median GA-RI was 0.50; reference interval: !0.25. Median BMI-adjusted amplitude was 37.6 µV; reference interval: 20-70 µV. The median ff-AR was 1.85; reference interval !1.08. Higher BMI was associated with a shorter meal-response duration (p=0.014).

**Conclusions:** This study provides normative reference intervals for BSGM spectral data that will be used in clinical practice to inform diagnostic interpretations of abnormal gastric function.

## Introduction

Chronic gastroduodenal symptoms such as nausea, vomiting, abdominal pain, early satiety, and excessive fullness affect approximately 10% of the global population.^1,2^ Despite a substantial healthcare and economic burden,^3^ progress toward defining specific disease mechanisms and patient subgroups has been slow.^4^ Current disease classifications depend on symptom-based criteria that overlap and fail to clearly delineate disease subgroups,^5,6^ while the role of gastric emptying testing is controversial due to debate regarding disease specificity, clinical utility, and instability over time.^5^ Novel diagnostic approaches are required to advance individualized therapy.^7^

Body surface gastric mapping (BSGM) is a new diagnostic test for evaluating gastric function.^8^ BSGM employs a dense grid of electrodes to non-invasively map gastric activity in high-resolution at the epigastrium, registering the myoelectrical activity that coordinates gastric motility and peristaltic activity through an increase in signal strength.^9,10^ BSGM represents a critical advance over traditional electrogastrography (EGG) approaches due to a combination of technical improvements that serve to maximally extract the weak underlying gastric signals while minimizing contamination by noise. These features include a high-resolution (HR) array of large area that accounts for gastric anatomical biodiversity while allowing signal summation from multiple sites, bioelectronics and signal processing systems specifically optimized for gastric electrophysiology, and a robust validated artifact detection and rejection scheme.^10–12^ Recent studies employing BSGM have revealed several disease signatures in adults and children suffering chronic nausea and vomiting syndromes, gastroparesis and functional dyspepsia, which correlated with symptoms.^9,11,13^ The first commercial medical device for performing BSGM has recently become available, with United States Food and Drug Administration clearance (Gastric Alimetry®, Alimetry, New Zealand), enabling clinical use with the goal of reducing diagnostic uncertainty.^10,11,14^

Standardised clinical application of a diagnostic medical physiology device requires a thorough understanding of normative metric intervals. For BSGM, two categories of metrics are recognized: i) *spectral metrics* encompassing myoelectrical frequency, amplitude, rhythm stability, and meal responses; and ii) *spatial metrics* characterising wave propagation dynamics.^9–11^ Spectral metrics are derived from a spectral plot, and represent a form of HR-EGG, achieving robust improvements over traditional EGG metrics.^8,10,12,15^ Spectral metrics are currently the most established form of analytics, with a recent BSGM study principally employing these to identify a distinct subgroup of patients with gastric neuromuscular abnormalities in nausea and vomiting syndromes.^11^

The aim of this study was to establish normative reference intervals for spectral BSGM metrics for Gastric Alimetry, encompassing the Principal Gastric Frequency, Gastric Alimetry Rhythm Index (GA-RI), BMI-adjusted amplitude, and the fed:fasted amplitude ratio.^15^ These intervals will be applied to guide clinical interpretation and provide clarity regarding biological variability in human gastric myoelectrical activity.

## Methods

This was an observational cohort study conducted in Auckland (New Zealand), Calgary (Canada), Louisville (KY, USA) and Western Sydney (Australia), (Ethical approvals references: AH1130, REB19-1925, 723369, H13541). All patients provided informed consent. The study is reported in accordance with the STROBE statement.^16^

### Healthy controls

Healthy subjects aged ≥18 years were recruited by local advertisement between 2021 and 2022. Screened subjects were exclueded if they had active gastrointestinal symptoms or diseases and regular medications affecting gastric motility or cannabis, or met Rome IV Critetia for a gastroduodenal disorder.^17^ Specific exclusion criteria related to BSGM, per the Gastric Alimetry Instructions For Use (IFU) were BMI >35 kg/m^2^, active abdominal wounds or abrasions, fragile skin, and allergies to adhesives. Pregnancy and previous upper GI surgeries were additional exclusion criteria.

### Study Protocol

The Gastric Alimetry System encompasses a HR stretchable electrode array (8×8 electrodes; 20 mm inter-electrode spacing; 196 cm^2^), a wearable Reader, an iPadOS App for set-up, anthropometric measurement-based array placement, and concurrent symptom logging during the test, and a cloud-based analytics and reporting platform.^10,11,14^ The standard Gastric Alimetry test protocol was followed.^10^ Participants fasted for >6 hrs and avoided caffeine and nicotine prior to testing. Array placement was preceded by shaving if necessary, and skin preparation (NuPrep; Weaver & Co, CO, USA). Recordings were performed for a fasting period of 30 minutes, followed by a 482 kCal meal consumed over 10 minutes and a 4-hr postprandial recording in order to capture a full gastric activity cycle. The meal consisted of Ensure (232 kcal, 250 mL; Abbott Nutrition, IL, USA) and an oatmeal energy bar (250 kcal, 5 g fat, 45 g carbohydrate, 10 g protein, 7 g fibre; Clif Bar & Company, CA, USA). Participants sat reclined in a chair and were asked to limit movement, talking, and sleeping, but were able to read, watch media, work on a mobile device, and mobilize for comfort breaks. Any subject reporting significant gastroduodenal symptoms during the test, via the validated Gastric Alimetry App,^14^ were excluded on the basis that they may have a subclinical disorder; transient symptoms of ‘excessive fullness” lasting ≤2.5 hours or minimal symptoms (≤2/10) were allowale.

### Data Analysis

Test quality was assessed based on impedance (<200 kΩ optimal; <500 kΩ acceptable; ≥500 kΩ poor) and proportion of the recording duration affected by artifacts (defined per the Gastric Alimetry Report Interpretation Guidelines as <20% optimal; <50% acceptable; >50% interpret with caution). Artifacts were automatically detected and rejected where possible using the validated Gastric Alimetry algorithm,^12^ and tests with artifacts >50% of the total testing duration were excluded from the reference interval calculations.

Spectral analysis of BSGM data is used to interpret the bioelectrical slow-waves that coordinate gastric motility, with gastric contractile activity also being registered as a power increase arising after a stimulus.^18,19^ Key physiological features of gastric myoelectrical activity motivated the development of spectral metrics used to establish the BSGM reference interval.^10,20–22^ These features and the corresponding metrics were recently described in detail by Schamberg et al,^15^ and are briefly outlined below, and depicted in **Figure 1**. Each metric is reported for the overall test, as well as for the preprandial period, postprandial period, and each 1-hour window postprandially. Metrics were not reported in time periods where over 50% of the data was removed by the Gastric Alimetry artifact detection and removal algorithm.^12^

**Figure 1.**
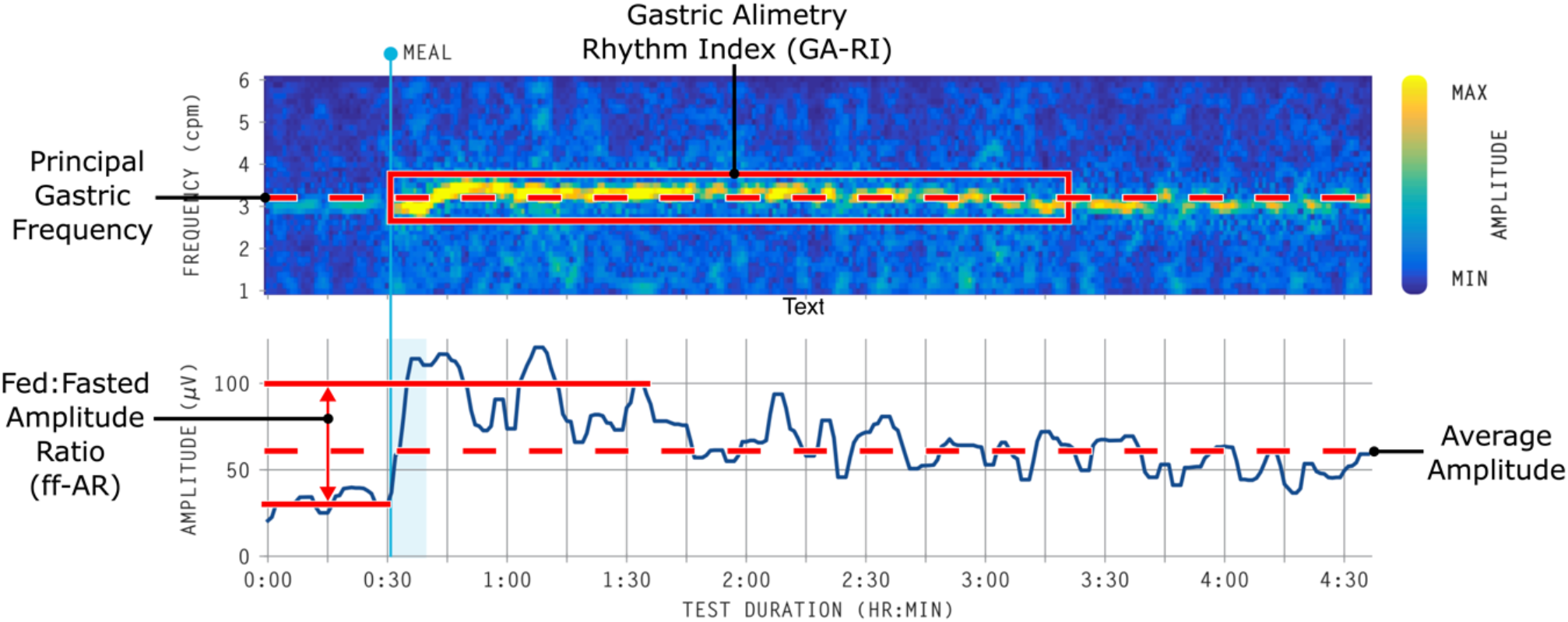
Example spectral plot for a single participant, with annotations of BSGM spectral metrics as recorded by the Gastric Alimetry System: Principal Gastric Frequency, Gastric Alimetry Rhythm Index (GA-RI), average amplitude, and fed:fasted amplitude ratio (ff-AR).

#### Amplitude

Amplitude (μV) reflects all active electrophysiological activity occurring within the gastric frequency range (i.e., both slow waves and smooth muscle contraction currents) under the region of the array and summated at the body surface.^23,24^ Amplitude is plotted as the maximum amplitude of the spectrum for each time point within the recorded duration, and the amplitude is reported as an average across time. The average amplitude is highly correlated with BMI in a non-linear manner,^8^ hence a BMI correction was applied to achieve a BMI-adjusted reference interval using a multiplicative regression model.^15^

#### Principal Gastric Frequency

The frequency (cycles per minute; cpm) of gastric slow waves has long been applied as a disease indicator for gastric myoelectrical activity.^20–22^ We previously reported that simply computing the maximum of the average spectrum did not reliably identify a gastric frequency consistent with visual assessment,^15^ so we developed a novel Principal Gastric Frequency metric, which identifies the gastric frequency based on maximizing the GA-RI (defined below).^15^ The Principal Gastric Frequency is not reported where the GA-RI does not identify a stable gastric slow wave, or for time periods with an average amplitude of <10 μV, which is the reliability threshold for the Gastric Alimetry system.

#### Gastric Alimetry Rhythm Index™ (GA-RI)™

‘Rhythm stability’ provides a measure of the extent to which the stomach is generating stable, organised, and consistent gastric slow waves. Measures of stability have previously been proposed to define discoordinated periods of gastric activity, such as instability coefficients of gastric frequency and amplitude/power.^20^ We previously reported that the use of instability coefficients may lead to incorrect assumptions about gastric stability,^15^ and therefore introduced GA-RI for BSGM, being a measure of the concentration of activity within the gastric frequency band relative to the residual spectrum over time.^15^ A high GA-RI occurs in cases where there is a distinct band of activity at a single frequency for an extended period, whereas a low GA-RI is observed in cases demonstrating a high degree of scatter in the spectral plot.^11^ GA-RI is not reported for time periods with an average amplitude of <10 μV. Additionally, we found that rhythm stability metrics are highly correlated with BMI.^15^ The GA-RI therefore includes an adjustment for BMI using a multplicative regression model, after which it is scaled to take on values ranging from zero (lowest stability) to one (highest stability).

#### Fed:Fasted Amplitude Ratio (ff-AR)

Many previous studies have reported that the amplitude of the cutaneous gastric signal typically increases following the consumption of a meal in healthy subjects.^20^ Calculating this metric to a short postprandial window, such as 45 mins to 1 hour leads to inaccurate ratio measurement, therefore, the BSGM ff-AR metric was defined as the ratio of the maximum 1-hour averaged postprandial amplitude (i.e. the average amplitude in either the 0-1 hour, 1-2 hour, 2-3 hour, or 3-4 hour postprandial window) to the pre-meal average amplitude.^15^ This measure is understood to primarily reflect the onset of gastric contractile activity in response to a meal.^19^ Unlike the other metrics, the ff-AR is only reported as an overall metric for each subject.

### Sample size

A cohort of 110 healthy controls were used to establish normative reference intervals for BSGM spectral metrics. This sample size is consistent with several studies developing normative reference intervals in gastrointestinal motility.^25,26^ Adequacy of the sample size was further assessed using bootstrap and cross-validation analyses (see <u>Statistical analysis</u> and **Supplementary Appendix**). To account for correlations between patient characteristics and BSGM metrics, the amplitude and stability metrics are adjusted for BMI as noted above, and all reported results were stratified by sex.

### Statistical analysis

Univariate associations between participant characteristics (age, sex, BMI, ethnicity) and spectral BSGM metrics (BMI-adjusted amplitude, Principal Gastric Frequency, GA-RI, ff-AR) were assessed with univariate regression models (age, BMI), t-test (sex), or the one-way ANOVA (ethnicity). Multivariate linear regression models were used for BSGM metrics with multiple significant univariate associations. For all regression analyses we report standardized coefficients (mean centred and standardised beta coefficients due to the different unit scales of variables in the regression models). Significantly skewed data (amplitude and ff-AR) were log-transformed prior to analysis. The median, 5^th^ and 95^th^ percentiles were estimated using the median-unbiased estimator,^27^ and were reported for each metric for the entire cohort and stratified by sex. 90% confidence intervals for the median, 5^th^ and 95^th^ percentiles were computed using a non-parametric bootstrap method with 1000 bootstrap samples. The normative reference intervals were then defined by rounding the 5^th^ percentile for one-sided reference intervals (ff-AR and GA-RI) and 5^th^/95^th^ percentiles for two-sided intervals (BMI-adjusted amplitude and Principal Gastric Frequency). Applicability of the reference interval to external subjects (i.e. those not included in the interval calculation) was assessed using a cross-validation analysis, wherein the complete cohort was split into five equal sized groups, with each group in turn being used to evaluate the fitness of reference interval and adjustment parameters computed using the remaining four groups (see **Supplementary Appendix**). All analyses were performed in Python v3.9.7 and R v.4.0.3 (R Foundation for Statistical Computing, Vienna, Austria).

## Results

BSGM using Gastric Alimetry was completed in 116 subjects. Of these, six subjects were excluded based on test quality whereby >50% of the recording was contaminated by artifacts. Of the remaining 110, the median age was 32 (IQR 24-50; range 18-73), the median BMI was 23.8 kg/m^2^ (IQR (21.4-26.9; range 16.6-35.0), 61 were female, seven were African/African-American, 17 were Asian, 73 were European/White, seven were Indian/Sri-Lankan, and six were other ethnicities. The average amplitude was >10 μV in all subjects. Two subjects did not have an identifiable Principal Gastric Frequency and were excluded from calculation of the Principal Gastric Frequency reference interval. One subject was excluded from the ff-AR reference interval due to having over 50% of the data removed from the preprandial window by the automated artifact removal algorithm (**Figure S1**).

### Associations between demographic characteristics and BSGM spectral variables

Age was not associated with any metric in univariate linear regression analyses and ethnicity was not associated with any metric in one-way ANOVA analyses (**Table S1**). BMI was weakly associated with ff-AR (coefficient=-0.22, p=0.023), and strongly associated with unadjusted/unscaled GA-RI (coefficient=-0.34, p<0.001). BMI was also strongly associated with amplitude on both univariate (coefficient=-0.55, p<0.001) and multivariate (coefficient=-0.54, p<0.001) analyses. Sex was associated with Principal Gastric Frequency (t-statistic=2.33, p=0.022) and amplitude in univariate (t-statistic=2.51, p=0.014) and multivariate (coefficient=0.21, p=0.007) analyses. Sex was not associated with GA-RI or ff-AR (p=0.32 and p=0.88). These results are summarised in **Table S1** and **Figure 2**. Based on these results, we report a BMI-adjusted amplitude and GA-RI, and all normative intervals for metrics were reported stratified by sex.

**Figure 2.**
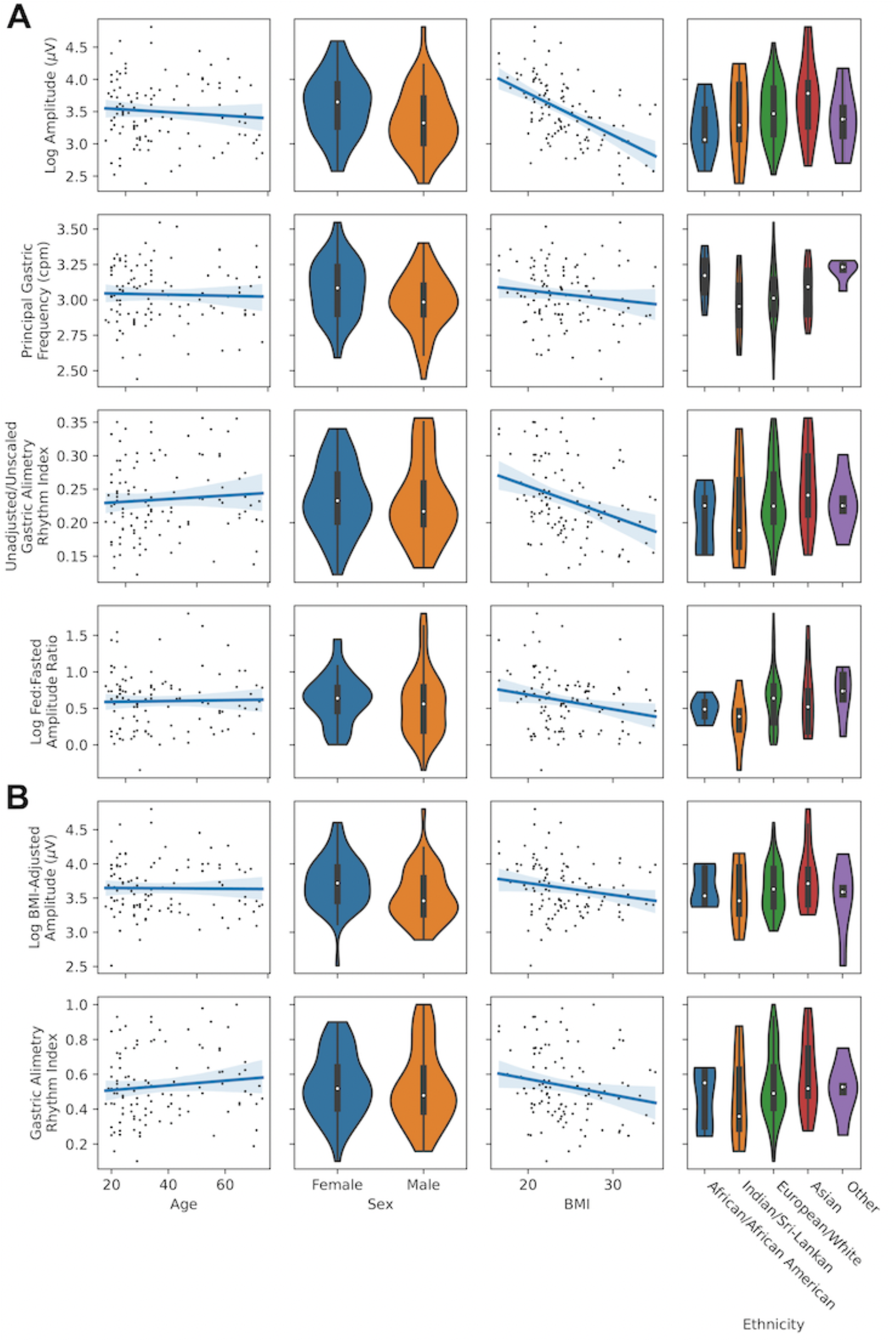
Association between patient demographic characteristics (age, sex, BMI, ethnicity) and BSGM metrics. A: Associations between demographic factors and unadjusted metrics (Log Amplitude, Principal Gastric Frequency, Unadjusted/Unscaled GA-RI, Log ff-AR); B: Association between demographic factors and BMI-adjusted metrics (BMI-adjusted amplitude, GA-RI).

The average spectrogram and BMI-adjusted amplitude curve for healthy controls is depicted in **Figure 3**. This demonstrates that a stable Principal Gastric Frequency, strong GA-RI, and a meal response are characteristic features of gastric myoelectrical function and define a normal BSGM spectral analysis. The Principal Gastric Frequency and BMI-adjusted amplitude were slightly lower in males than females (median 2.98 cpm and 31.81 μV in males vs 3.09 cpm and 41.22 μV in females; p=0.022 and p=0.033 respectively; **Figure 3B and 3C)**. The Principal Gastric Frequency and BMI-adjusted amplitude were lower on average in the preprandial period than the postprandial period (median 2.94 cpm and 26.57 μV preprandial vs 3.05 cpm and 39.38 μV postprandial; p=0.001 and p<0.001 respectively).

**Figure 3.**
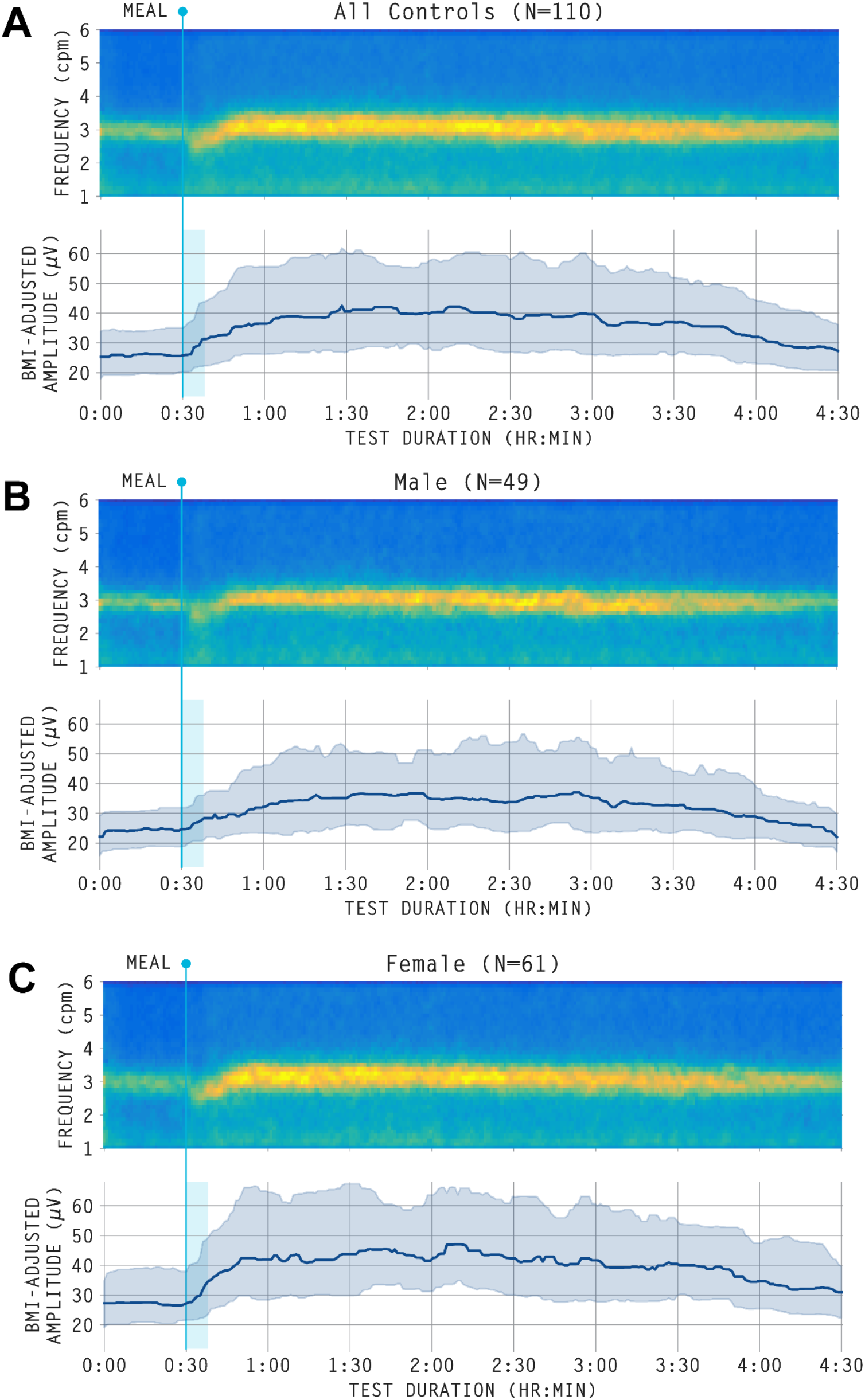
Averaged spectrogram (top) and median BMI-adjusted amplitude curve with shaded interquartile range (bottom). A: whole cohort; B: males; C: females. All amplitude curves are displayed after applying a median filter for visual clarity.

### Normative reference intervals for BSGM metrics

The median, 5^th^ percentile, 95^th^ percentile and associated 90% bootstrap confidence intervals for each metric, computed overall and stratified by sex, are summarised in **Table 1**. Recommended reference intervals for identifying abnormal gastric activity from BSGM spectral metrics are given in **Table 2**. The median and IQR for BMI-adjusted amplitude, Principal Gastric Frequency, and GA-RI for the preprandial period, postprandial period, and each 1-hour window postprandially are reported in **Table S2**. Results of the cross-validation analysis are provided in the **Supplementary Appendix** and **Table S3**. Representative spectrograms of cases within this healthy control cohort that fell outside the reference interval for each metric are depicted in **Figure S2**.

**Table 1:** Normative healthy control data reporting median, 5th and 95th percentiles with associated 90% confidence intervals for the overall cohort and stratified by sex for BMI-adjusted amplitude, Principal Gastric Frequency, GA-RI, and ff-AR.

| Metric | Stratification | 5th Percentile (90% CI) | Median (90% CI) | 95th Percentile (90% CI) |
| --- | --- | --- | --- | --- |
| BMI-Adjusted Amplitude (μV) | Overall | 21.29 (20.46 - 24.40) | 37.64 (33.34 - 40.03) | 71.89 (63.06 - 97.16) |
|  | Male | 20.52 (17.98 - 22.05) | 31.81 (29.49 - 37.53) | 61.63 (54.38 - 121.61) |
|  | Female | 26.30 (16.44 - 27.95) | 41.22 (37.76 - 44.51) | 81.88 (63.65 - 98.67) |
| Principal Gastric Frequency (cpm) | Overall | 2.65 (2.61 - 2.76) | 3.04 (3.00 - 3.09) | 3.35 (3.31 - 3.40) |
|  | Male | 2.62 (2.44 - 2.72) | 2.98 (2.94 - 3.05) | 3.30 (3.23 - 3.40) |
|  | Female | 2.75 (2.61 - 2.84) | 3.08 (3.03 - 3.15) | 3.37 (3.32 - 3.54) |
| Gastric Alimetry Rhythm Index | Overall | 0.25 (0.19 - 0.28) | 0.50 (0.48 - 0.54) | 0.90 (0.86 - 0.94) |
|  | Male | 0.24 (0.16 - 0.27) | 0.48 (0.43 - 0.53) | 0.94 (0.87 - 1.00) |
|  | Female | 0.25 (0.16 - 0.32) | 0.52 (0.49 - 0.56) | 0.86 (0.78 - 0.89) |
| Fed:Fasted Amplitude Ratio | Overall | 1.08 (1.01 - 1.12) | 1.85 (1.75 - 1.95) | 4.13 (2.90 - 4.68) |
|  | Male | 1.06 (0.71 - 1.13) | 1.75 (1.43 - 1.92) | 4.76 (2.90 - 6.04) |
|  | Female | 1.09 (1.01 - 1.17) | 1.89 (1.79 - 2.05) | 3.57 (2.72 - 4.23) |

**Table 2:** Normative reference intervals for BMI-adjusted amplitude, Principal Gastric Frequency, GA-RI, and ff-AR

| Metric | Lower | Upper |
| --- | --- | --- |
| BMI-Adjusted Amplitude (μV) | 20 | 70 |
| Principal Gastric Frequency (cpm) | 2.65 | 3.35 |
| Gastric Alimetry Rhythm Index | 0.25 | - |
| Fed:Fasted Amplitude Ratio | 1.08 | - |

### Additional features of gastric function

#### High fasting baseline

A significant subset of controls presented with a high fasting baseline amplitude. This is illustrated in **Figure 4**, which shows the average spectrograms and amplitude curves for cases with low ff-AR (≤1.5) and high ff-AR (>1.5) (refer also individual case examples in **Figure S5)**. During the preprandial period, the BMI-adjusted amplitude was 28.19 μV (IQR 25.34-33.64) for low ff-AR group compared with 25.21 μV (IQR 19.75-31.47; p=0.009) for the high ff-AR group (**Figure 4.C**). Therefore, the lack of an apparent meal response in some cases can be attributed in part to higher baseline amplitude before the meal (as opposed to the lack of activity following meal consumption).

**Figure 4:**
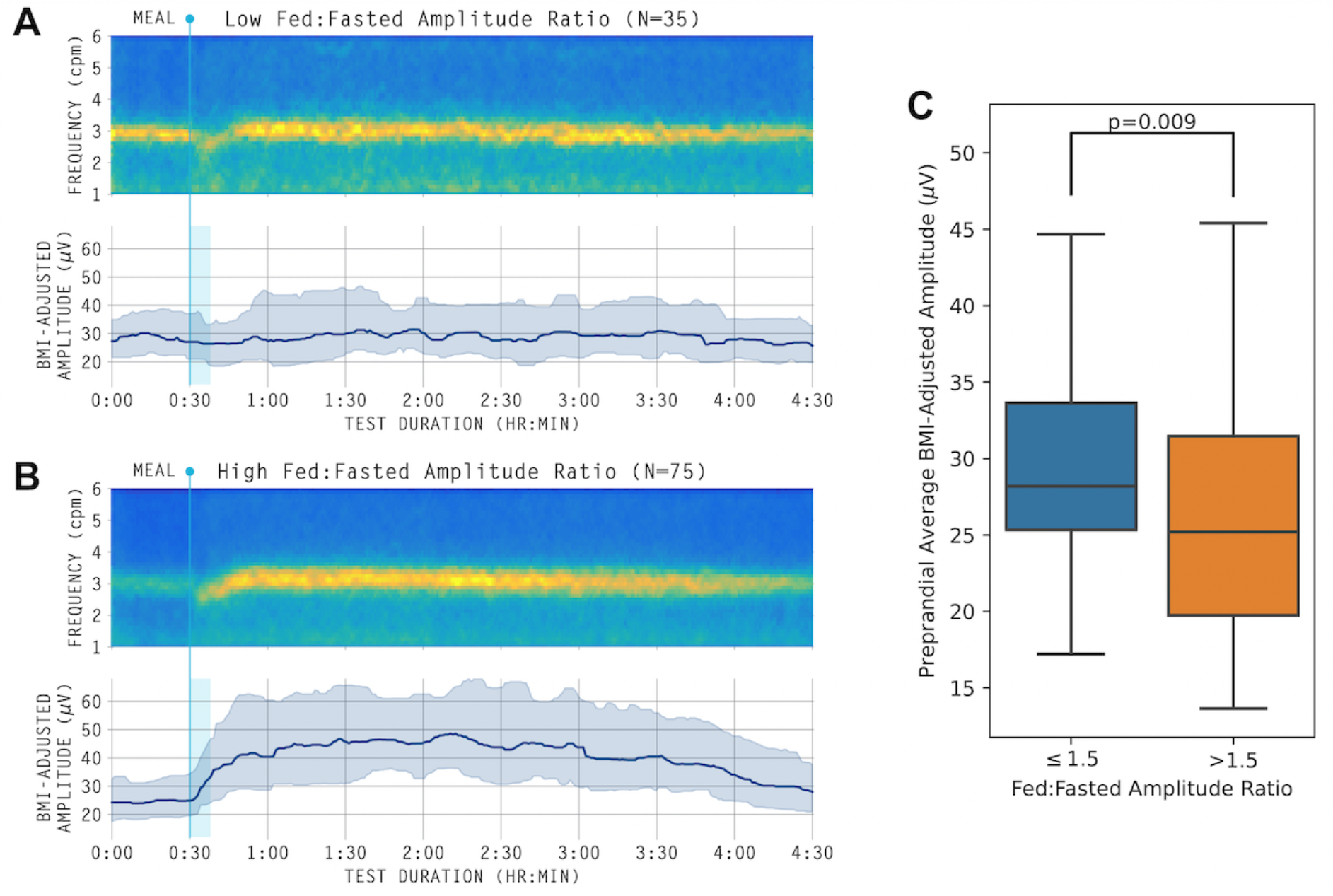
Averaged spectrogram and median BMI-adjusted amplitude curve with shaded interquartile range for low and high ff-AR cases. A: Low ff-AR (≥1.5); B: High ff-AR (>1.5); C: Distribution of preprandial average amplitude for the two groups.

#### Differences in the meal response duration based on BMI

To explore the association between BMI and spectral BSGM data, separate average spectrograms were plotted for those with BMI <25 and ≥25 with unadjusted and adjusted amplitude curves (**Figure 6**). Average spectrograms were similar between BMI groups, however, it was visually apparent that subjects with higher BMIs within the studied range may have a shorter meal response duration, as shown by earlier down-trending of amplitude curves on the summary spectral plots. This visual trend was supported by a significant difference in the time of the peak amplitude (low BMI median 2.12 hours after meal completion (IQR 0.98-2.72) vs high BMI median 0.97 hours after meal completion (IQR 0.28-1.76; p=0.014; **Figure 5**). This preliminary analysis indicates that the timescale of the meal response is more rapid in subjects with high BMI than those with low BMI.

**Figure 5.**
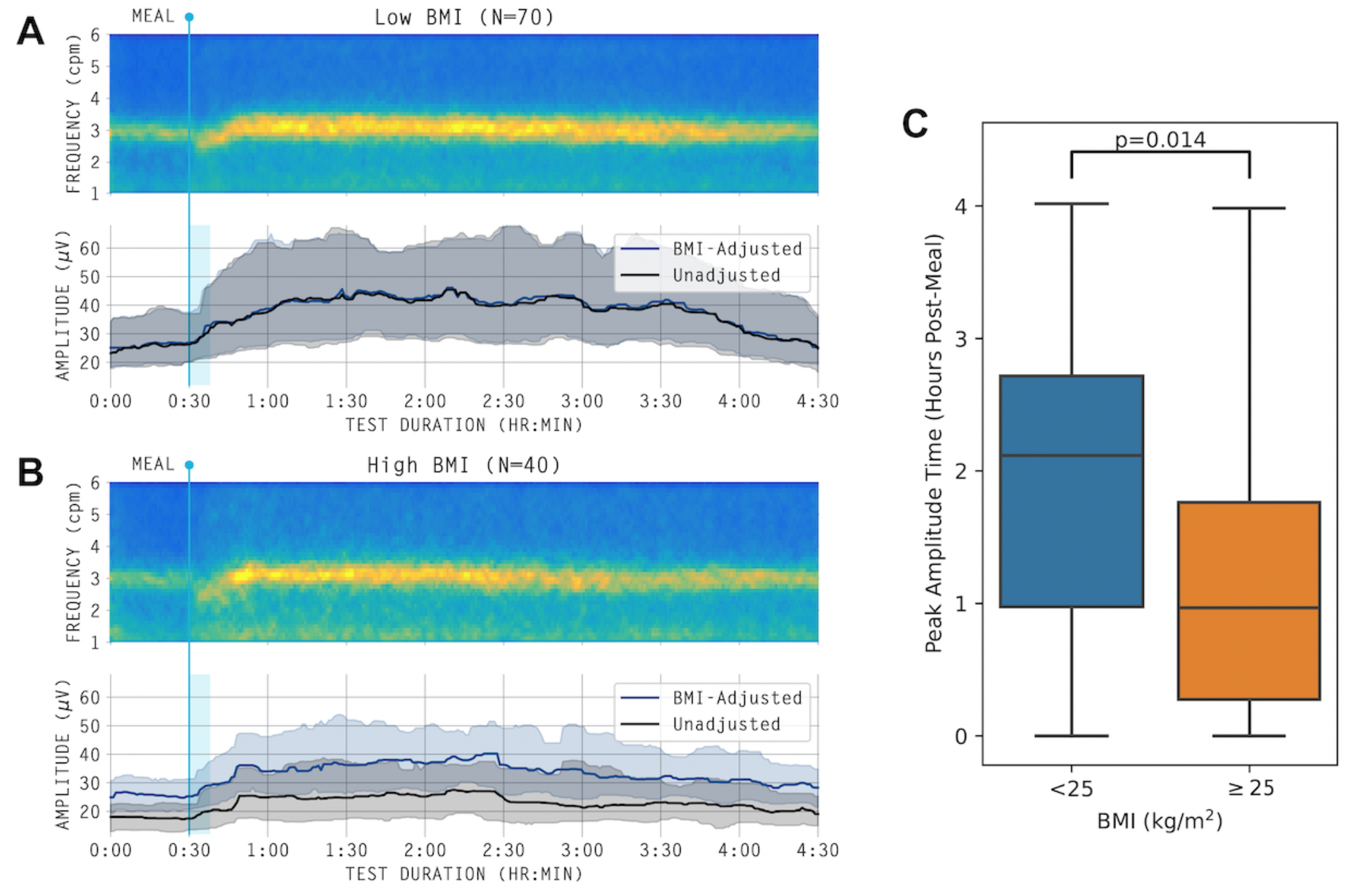
Averaged spectrogram and median amplitude curves (BMI-adjusted and unadjusted) with shaded interquartile range for low and high BMI cases. A: Low BMI (<25 kg/m^2^); B: High BMI (≥25 kg/m^2^); C: Times of peak postprandial amplitude for the two groups.

## Discussion

This study defines normative reference intervals for BSGM spectral metrics when recorded using the Gastric Alimetry system, and has characterized and clarified normal gastric electrophysiology in a healthy control population. These data will be instrumental in guiding interpretations of BSGM test data in clinical practice.

Although systematic reviews of EGG have shown that gastric myoelectrical abnormalities are prevalent in patients with nausea and vomiting disorders,^28^ functional dyspepsia,^29^ gastro-esophageal reflux,^30^ as well as in paediatric and post-operative disorders,^31^ the test has not achieved common clinical acceptance due to concerns surrounding validity and utility.^8,32,33^ BSGM represents a more reliable diagnostic tool for evaluating gastric function,^8,10^ while remaining non-invasive. Early studies have confirmed improved symptom correlations and the ability to separate phenotypes at the individual patient level.^9,11^ The optimized set of spectral metrics with reference intervals provided here will be fundamental in advancing the clinical application of BSGM, while promoting methodological and data standardization in research.

A recent study applying BSGM in patients with nausea/vomiting syndromes including gastroparesis demonstrated that nearly a third of cases featured abnormal spectrograms indicative of gastric neuromuscular dysfunction.^11^ This phenotype was characterized by low amplitude or absent gastric frequency bands, poor rhythm stability, and limited meal responses, allowing identification of a distinct subgroup despite non-specific symptom profiles. However, this previous subgroup determination relied on subjective categorization by a consensus panel. The new GA-RI metric was informed by these findings, and when GA-RI falls below the 5^th^ percentile of the reference interval, the spectrograms visually match those from the gastric dysfunction subgroup.^11^ The GA-RI and its associated reference interval therefore present a new objective tool for routinely differentiating groups of patients with gastric neuromuscular abnormalities.

Although frequency has been the most prominent metric in the EGG literature, there has previously been no consensus on what constitutes normal, with intervals such as 2-4 cpm and 2.5-3.7 cpm commonly cited.^20,34^ By focusing on the frequency associated with stable and persistent gastric activity, we find that the true reference interval is considerably narrower at 2.65-3.35 cpm, which is also closer to that verified by direct serosal recordings.^22^ This refinement reflects the higher signal-to-noise ratio of BSGM, as well as the pitfall that legacy EGG metrics could conflate scattered/unstable gastric activity or noise with low frequency gastric activity.^15,35^ As such, the metrics used for establishing the reference intervals here were intentionally structured to independently capture the gastric frequency and the stability of the activity at that frequency. This concept is illustrated in **Figure S4**, which depicts a challenging spectrogram for a subject with a normal gastric frequency but low GA-RI. The new metrics are therefore able to distinguish between gastric contractions that occur at an abnormally low-frequency (e.g. **Figure S2.A**) and normal frequency activity occurring simultaneously with uncoordinated, low frequency activity which could be related to noise (e.g., simultaneous colonic activity).^36^ In addition, our data reveal a significant sex difference in gastric frequency and BMI-adjusted amplitude, with both being slightly higher in females, which is a new finding not seen in previous EGG studies.^37^ However, the magnitude of this difference was sufficiently small as to not warrant separation of the Principal Gastric Frequency reference interval by sex, and it is unlikely to have clinical relevance in test interpretation.

Amplitude has been of longstanding interest in the EGG literature, however, amplitude data previously was not validated for clinical applications due to the lack of a reference interval and substantial variability between patients due to diminution with abdominal adiposity,^38^ as well as intra-study variability following excitatory stimuli.^21^ These problems were overcome in this study by developing a BMI-adjusted amplitude reference interval that considers an entire 4-hr meal response cycle, thereby accounting for biological variability in meal response time-curves.^10^ We also elected to include both a low and high amplitude cut-off, recognising that low-amplitude activity may be a feature of gastric neuromuscular dysfunction,^11^ while high amplitude activity has been recognized to have diagnostic utility in relation to gastric outlet obstruction.^39^

Previous work has also demonstrated that an amplitude increase in response to a meal is a normal feature of cutaneous gastric recordings, with ff-AR metrics previously commonly employed in EGG studies, albeit over substantially shorter time periods.^10,20–22^ However, we noticed that many healthy controls demonstrate high fasting baseline amplitude at the gastric frequency, such that the perceived meal response is diminished or absent. This finding could be due to cephalic-phase gastric activity,^40^ migrating motor complex activity,^41^ or circadian variations.^42^ While the average spectrograms and median amplitude curves in **Figure 3** confirm a typical meal response at the cohort level (median ff-AR: 1.85), further evaluations of meal response dynamics will be necessary before ff-AR alone can offer clinical utility as a disease-discriminating metric. Nevertheless, its absence in combination with other spectral abnormalities is supportive of gastric neuromuscular dysfunction.^11^

The link between gastric motility and BMI is conflicting in the literature, but it has been postulated that differences in the gastric meal responses and emptying rates may play an etiologic role in obesity, predominantly through modulation of satiety.^43^ The shorter meal response observed in higher BMI individuals in this study was therefore an important new finding, because it may offer a non-invasive biomarker for therapeutics aiming to slow gastric function in the treatment of obesity and diabetes (e.g. GLP-1 receptor agonists).^44^ It remains unclear how the different meal response profiles identified here relate to gastric emptying, but this could be clarified in future by performing simultaneous BSGM and gastric emptying tests across a wide BMI range. Previous EGG studies have also suggested that there may be differences in gastric myoelectrical activity and autonomic responses between obese and lean individuals in response to varying meal compositions,^45^ which was not a focus of the present study. An in-depth analysis of the meal response curves and their relationship to BMI will be the subject of future research.

This study marks the largest collection of BSGM data on healthy controls, and care was taken to include a representative range of sex, age, ethnicity, and BMI to support broad clinical utility. The use of non-parametric bootstrap methods for computing confidence intervals provides an estimate of the extent to which we can expect to see variability in the reference intervals of these metrics as more data becomes available. Furthermore, the cross-validation analysis (see **Supplementary Appendix**) provides an estimate of the percentage of healthy controls from an independent cohort we would expect to fall outside of the reference interval for each of the metrics. Both the bootstrap and cross-validation analyses indicate that the spectral reference interval in its current form is validated for understanding and assessing normality of BSGM spectral metrics.

Nevertheless, some limitations to this work are acknowledged. The reference interval specifications are fundamentally dependent on the recording modality, and these data are therefore only relevant to tests performed with the Gastric Alimetry system. While this is the only BSGM system commercially available at this time, other systems have been used in research.^13,46^ Tests with >50% artifact were excluded from the reference interval calculations, and repeat tests may therefore be needed clinically where excessive artifacts occur, or interpretations made with caution. Substitute meals for patients with specific dietary requirements (e.g. gluten-free, diabetic) are expected to be inconsequential if similar caloric loads and nutritional content are employed, however further research is required to understand whether the reference intervals maintain their validity under shorter time periods, reduced caloric loads, or different test meals. In addition, while the associations of BMI with amplitude and GA-RI were accounted for by adjustment, other demographic associations were noted, specifically significant correlations between sex and amplitude, sex and Principal Gastric Frequency, and BMI and ff-AR. Given that the relationship between BMI and amplitude yields a clearer physiologic interpretation than that of sex and amplitude, and that there was no correlation between sex and BMI-adjusted amplitude, we hypothesize that sex/amplitude correlation reflects the correlation between sex and BMI. As the correlation between BMI and ff-AR was very weak, we did not perform an adjustment. Ongoing separate work is addressing intra-individual reproducibility of BSGM data. As the applications of BSGM expand, it is expected that further reference intervals will become available, and that further refinements to the understanding of normal gastric function will be required. In particular, a spatial reference interval is in development to provide wave direction and stability metrics, which have been shown to correlate with symptoms in functional dyspepsia and nausea and vomiting disorders, and these would further expand the clinical utility of BSGM.^9,11,13^

In conclusion, this study presents reference intervals for BSGM spectral data, incorporating novel and updated metrics. These data characterise normal human gastric electrophysiology at an unprecedented level of accuracy and will be essential to guide interpretations of Gastric Alimetry test data in clinical practice.

## Data Availability

Data are available on request from the corresponding author.

## Acknowledgements

We thank the volunteers who participated in this research and our clinical coordinating teams, Gen Johnston and India Wallace in Auckland, Lynn Wilsack and Renata Rehak in Calgary, and Abigail Stocker, Ben Rogers, Prateek Mathur, and Sara Elnour in Louisville.

---

**Figure S1:**
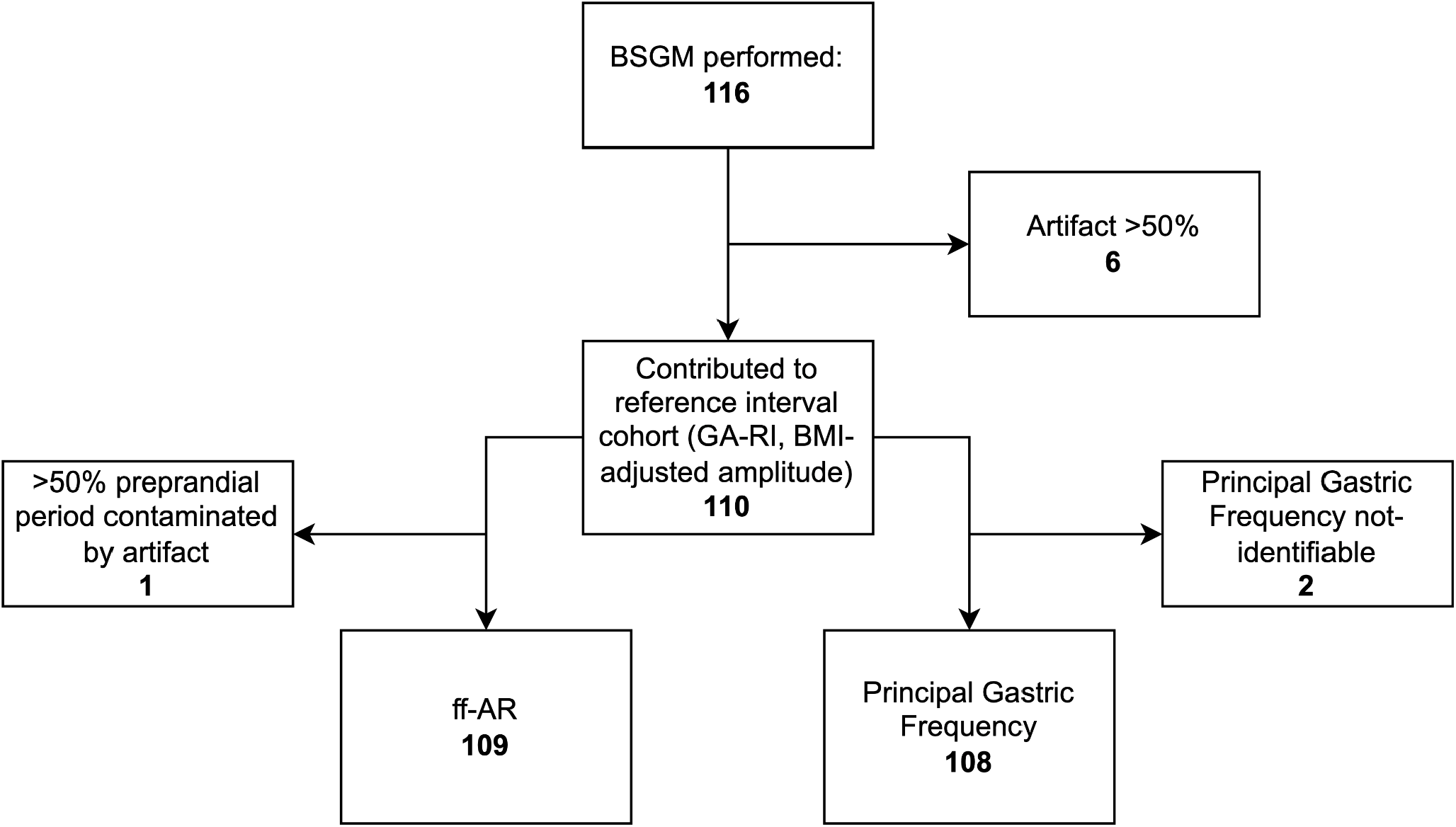
Patient flow diagram of subject exclusions with reasons.

**Figure S2:**
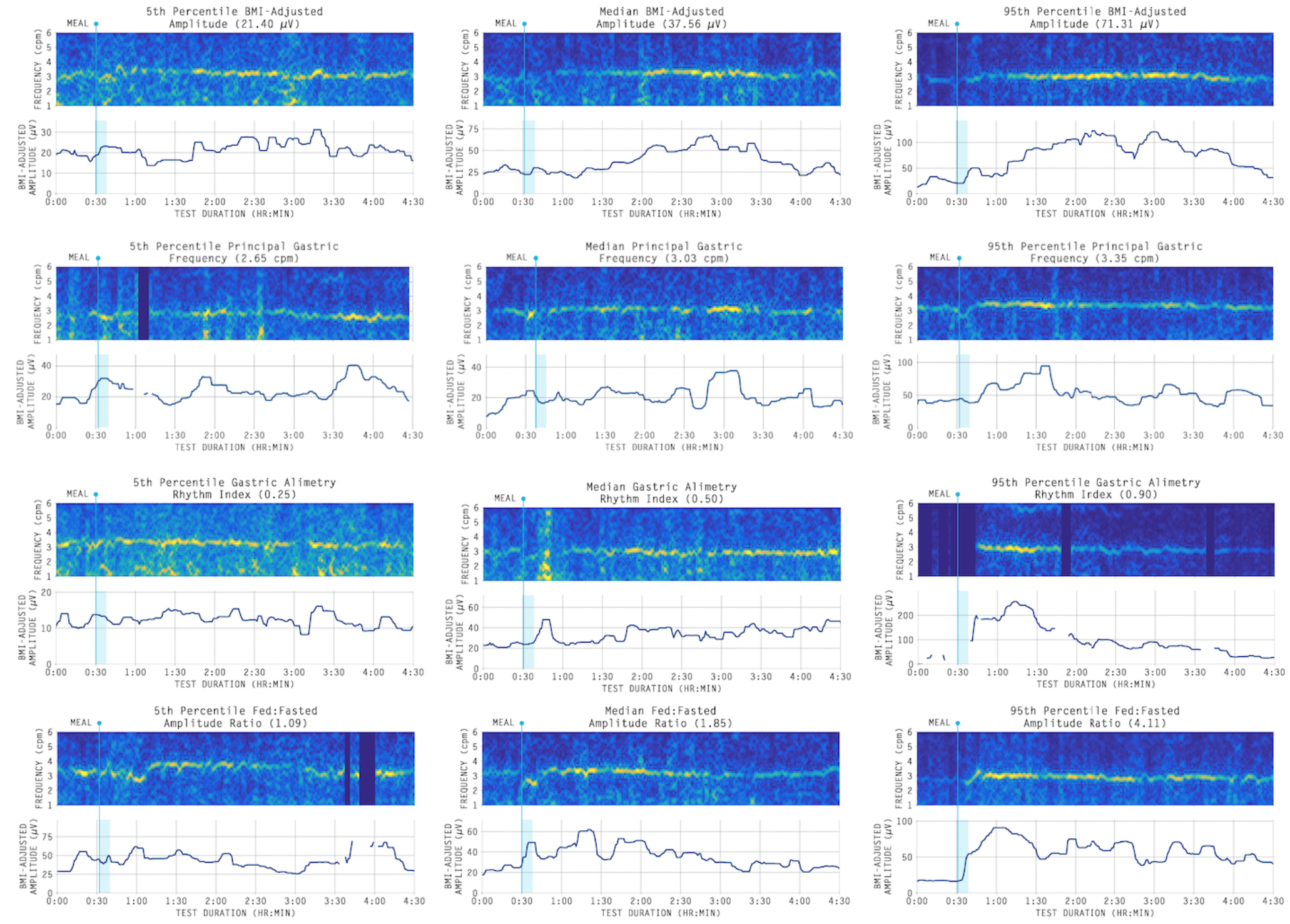
Spectrograms associated with the 5th, median, and 95th percentile for BMI-adjusted amplitude, Principal Gastric Frequency, GA-RI, and ff-AR.

**Figure S3.**
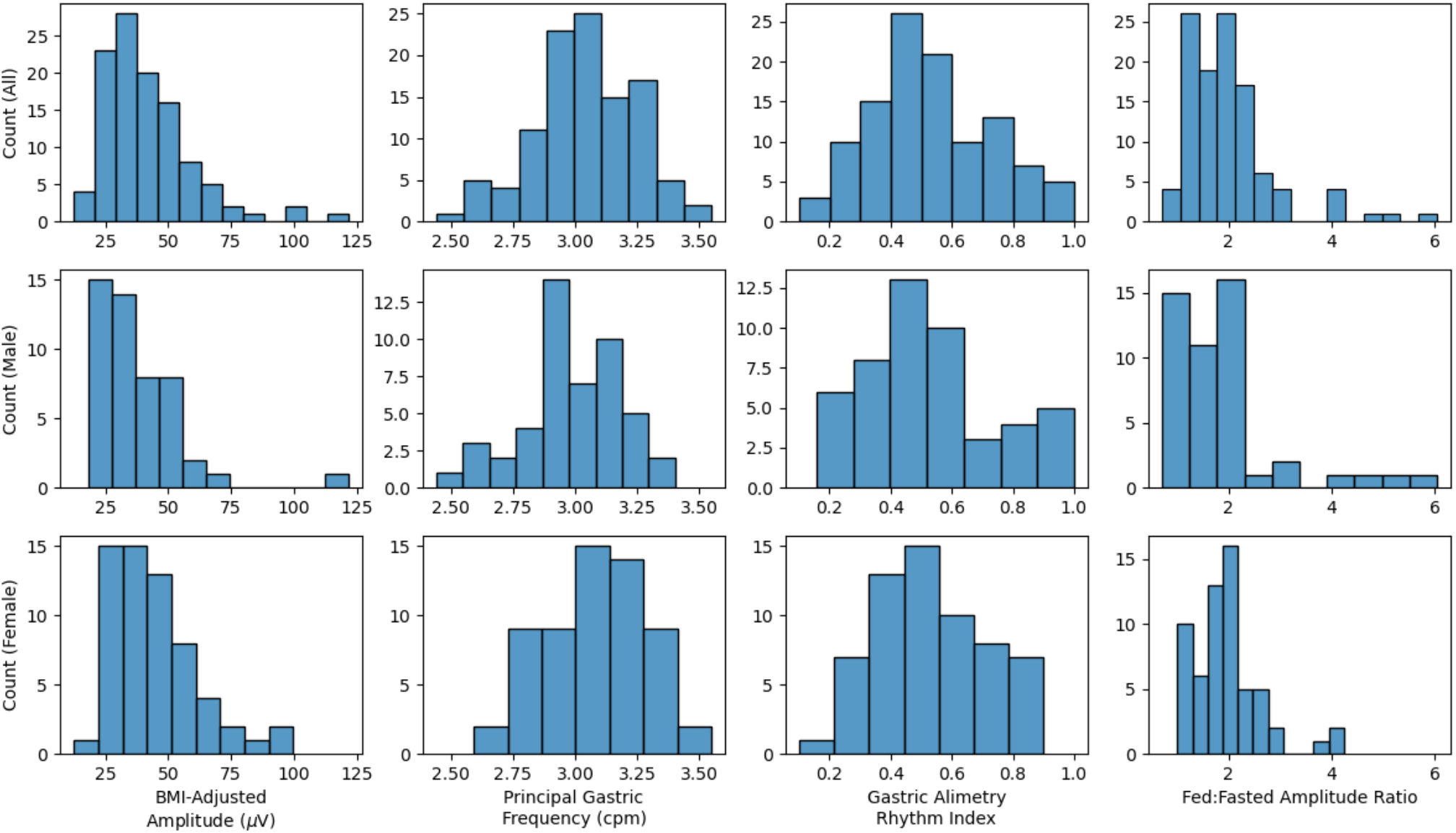
Histograms for BMI-adjusted amplitude, Principal Gastric Frequency, GA-RI, and ff-AR for the entire cohort and stratified by sex.

**Figure S4.**
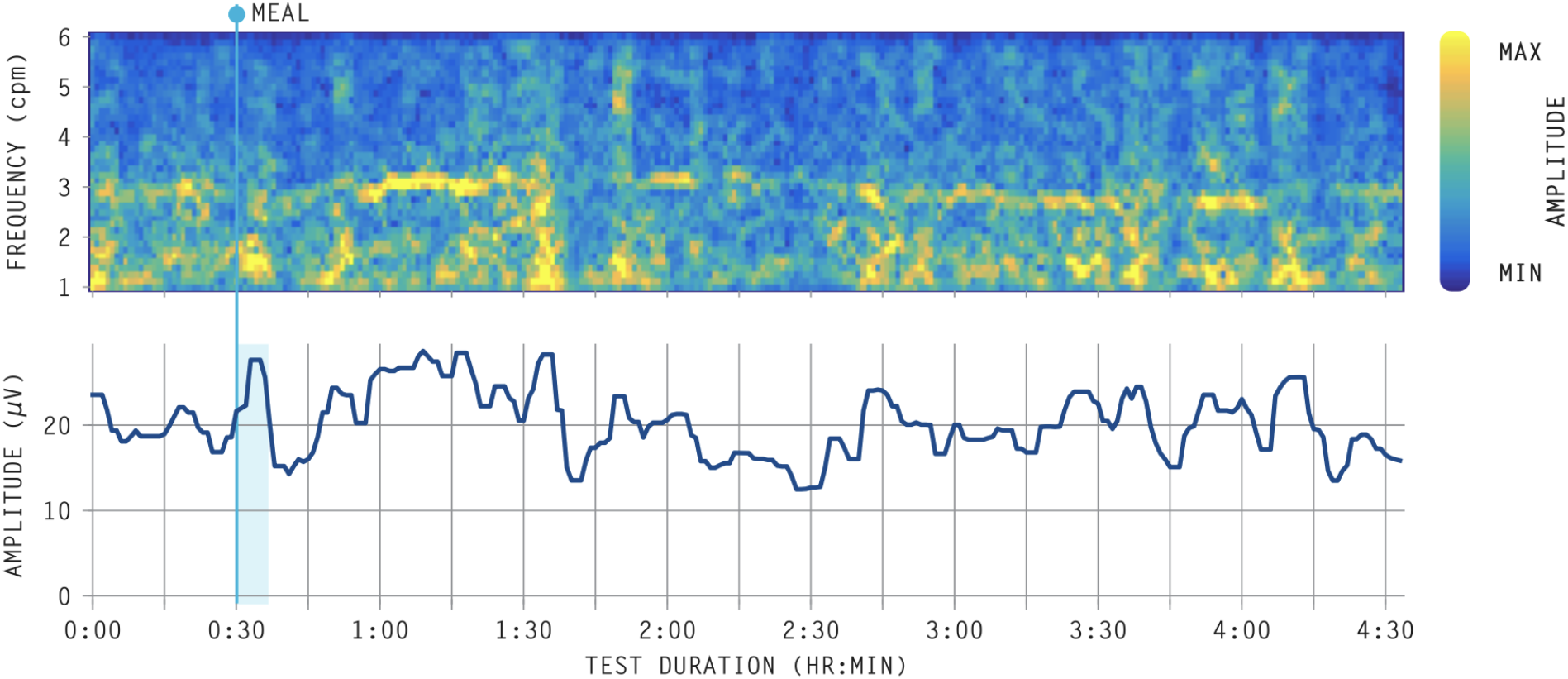
Spectrogram for a control with a normal but highly unstable spectral plot. Despite having a distinct band of activity near 3 cpm, traditional methods for identifying the gastric frequency yield an estimate of 1.1 cpm due to the high amplitude of the sporadic low frequency activity. By contrast, the Principal Gastric Frequency metric developed here is reported at 2.9 cpm, and the low frequency activity is instead captured by the GA-RI of 0.19, which is outside of the reference interval.

**Figure S5:**
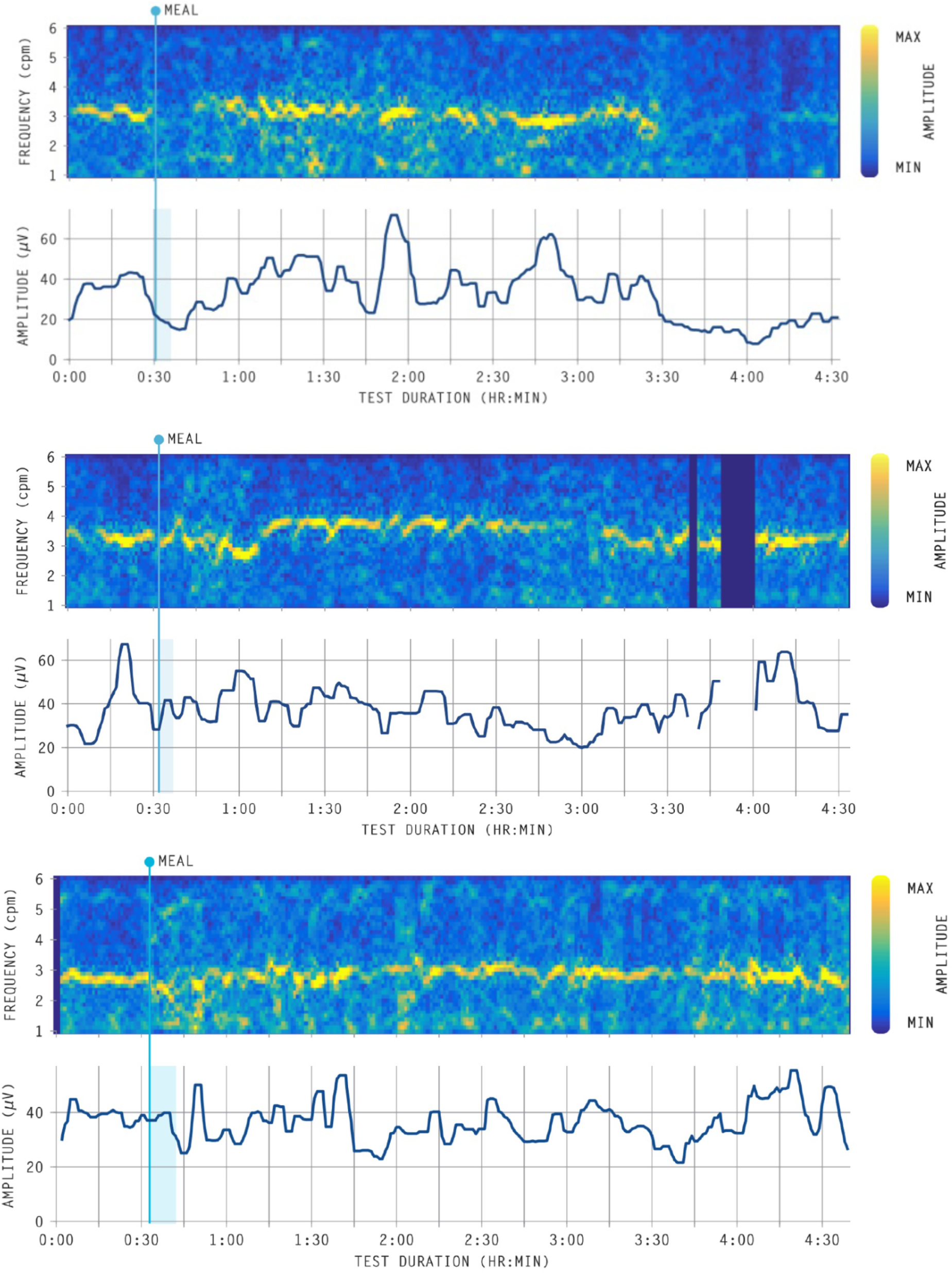
Examples (n = 3) of spectral and amplitude plot of a low meal response participants confounded by a high-fasting baseline amplitude.

**Table S1:** Univariate and multivariate models of the impact of patient characteristics (age, sex, BMI, and ethnicity) on metrics (Log Amplitude, Principal Gastric Frequency, Unadjusted/Unscaled GA-RI, Log ff-AR). The test statistic for the univariate models is either the standardised regression coefficient with 90% CI (age, BMI), t-statistic (sex), or F-statistic (ethnicity). The sex variable is referenced to male in the multivariate regression model.

| Metric | Characteristic | Univariate |  | Multivariate |  |
| --- | --- | --- | --- | --- | --- |
|  |  | Test Statistic | p | Standardised Coefficient (90% CI) | p |
| Log Amplitude | Age | -0.09<br>(-0.28 - 0.10) | 0.366 |  |  |
|  | Sex | 2.51 | 0.014 | 0.21 (0.06 - 0.37) | 0.007 |
|  | BMI | -0.55<br>(-0.71 - -0.39) | <0.001 | -0.54<br>(-0.69 - -0.38) | <0.001 |
|  | Ethnicity | 1.18 | 0.325 |  |  |
| Principal Gastric Frequency | Age | -0.03<br>(-0.22 - 0.16) | 0.748 |  |  |
|  | Sex | 2.33 | 0.022 |  |  |
|  | BMI | -0.13<br>(-0.32 - 0.06) | 0.179 |  |  |
|  | Ethnicity | 2.19 | 0.076 |  |  |
| Unadjusted/<br>Unscaled<br>Gastric Alimetry<br>Rhythm Index | Age | 0.07 (-0.12 - 0.26) | 0.437 |  |  |
|  | Sex | 0.32 | 0.748 |  |  |
|  | BMI | -0.34<br>(-0.51 - -0.16) | <0.001 |  |  |
|  | Ethnicity | 1.16 | 0.335 |  |  |
| Log Fed:Fasted<br>Amplitude Ratio | Age | 0.02 (-0.17 - 0.21) | 0.81 |  |  |
|  | Sex | 0.89 | 0.377 |  |  |
|  | BMI | -0.22<br>(-0.40 - -0.03) | 0.023 |  |  |
|  | Ethnicity | 1.25 | 0.293 |  |  |

**Table S2:** Median (IQR) metric values for specific time periods of each test

| Metric | Preprandial | Postprandial | 0-1hr<br>Postprandial | 1-2hr<br>Postprandial | 2-3hr<br>Postprandial | 3-4hr<br>Postprandial |
| --- | --- | --- | --- | --- | --- | --- |
| BMI-Adjusted<br>Amplitude ($\mu\text{V}$) | 26.58 (20.86 -<br>32.80) | 39.38 (30.33 -<br>53.21) | 37.74 (30.25 -<br>54.24) | 41.98 (30.51 -<br>58.04) | 37.59 (29.06 -<br>54.66) | 30.92 (26.02 -<br>43.40) |
| Principal<br>Gastric<br>Frequency<br>(cpm) | 2.94 (2.80 -<br>3.11) | 3.05 (2.90 -<br>3.19) | 3.10 (2.97 -<br>3.24) | 3.07 (2.94 -<br>3.26) | 2.96 (2.81 -<br>3.13) | 2.96 (2.83 -<br>3.10) |
| Gastric<br>Alimetry<br>Rhythm Index | 0.43 (0.29 -<br>0.62) | 0.51 (0.40 -<br>0.69) | 0.49 (0.31 -<br>0.69) | 0.55 (0.42 -<br>0.76) | 0.55 (0.43 -<br>0.72) | 0.52 (0.36 -<br>0.68) |

**Table S3:** Results of the cross-validation validation analysis. Each cell represents the percentage of subjects in that fold (n=22) falling outside a reference interval computed using the remaining four folds (n=88).

| Fold<br>Number | Low BMI-Adjusted<br>Amplitude | High BMI-Adjusted<br>Amplitude | Low<br>Principal<br>Gastric<br>Frequency | High<br>Principal<br>Gastric<br>Frequency | Low<br>Gastric<br>Alimetry<br>Rhythm<br>IndexIndex | Low<br>Fed:Fasted<br>Amplitude<br>Ratio | Mean<br>(Std. Dev.) |
| --- | --- | --- | --- | --- | --- | --- | --- |
| 1 | 0 | 4.55 | 0 | 0 | 9.09 | 0 | 2.27 (3.47) |
| 2 | 9.09 | 0 | 18.18 | 0 | 13.64 | 13.64 | 9.09 (6.94) |
| 3 | 4.55 | 9.09 | 0 | 0 | 4.55 | 0 | 3.03 (3.39) |
| 4 | 9.09 | 9.09 | 9.09 | 9.09 | 0 | 4.55 | 6.82 (3.47) |
| 5 | 0 | 0 | 0 | 13.64 | 0 | 4.55 | 3.03 (5.03) |
| Mean<br>(Std. Dev.) | 4.55 (4.07) | 4.55 (4.07) | 5.45 (7.27) | 4.55 (5.75) | 5.46 (5.30) | 4.55 (4.98) | 4.85 (5.37) |

## Supplementary Appendix: Methods

### Cross-validation analysis

Cross-validation is a commonly used technique for evaluating how well a statistical model will generalize to a set of data that was not used in the model fitting procedure. In the context of normative reference intervals, it is desirable to show that an independent sample of healthy controls will have the same percentage of metrics falling outside of the reference interval. To assess this in the proposed reference intervals, we performed a 5-fold cross-validation, wherein the cohort was randomly partitioned into five groups of size n=22 and each group was used to evaluate the fitness of a reference interval computed using the remaining four groups (i.e. using a cohort of size 88). For each of the five groups, we calculated the percentage of controls with metrics falling out of the reference interval. The results, shown in **Table S3**, suggest that we can expect approximately 5% of subjects to fall outside of the reference interval for each metric (10% for two-sided intervals) when analysing an independent sample of healthy controls.

